# Spatially Constrained Gene Regulation Identifies Key Genetic Contributions of Preeclampsia, Hypertension and Proteinuria

**DOI:** 10.1101/2022.05.13.22275064

**Authors:** Genevieve Boom, Justin M. O’Sullivan, William Schierding

## Abstract

Preeclampsia (PE) is a relatively common but severe pregnancy disorder that is characterized by hypertension (HTN) and either proteinuria (PRO), or other organ damage. There are very limited effective treatments for PE, and it is associated with substantial effects on the health of both the mother and fetus. 25 genetic variants have been associated (p < 1 × 10^−6^) with PE in the latest genome-wide association studies (GWAS). By overlapping the regulatory impacts of the PE-associated genetic variants with the regulatory impacts of HTN- and PRO-associated genetic variants, we were able to identify shared functional impacts between PE and HTN. We identified significant variation of the mean LOUEF scores for genes targeted by *cis*- and *trans*-acting eQTLs, consistent with an enrichment for regulatory interactions with target genes intolerant to loss-of-function mutations. Signaling pathways were enriched within the protein-protein interaction networks (PPINs) that were constructed using proteins encoded by the eQTL targeted genes. Finally, tissue-specific analyses of eQTLs specific to whole blood and arteries detected dysregulation of PE-relevant regulatory pathways. Collectively, these results are consistent with a model in which predisposition to HTN and PRO lays a molecular groundwork toward risk for PE pathogenesis. These findings inform on possible therapeutic targets for future studies.

## Introduction

Preeclampsia (PE) is a severe disease that affects 2-8% of all pregnancies, impacting pregnant women who were normotensive prior to pregnancy.^1^ The onset of PE typically occurs after 20 weeks of gestation, and PE can be categorized into subtypes according to the age of onset: early (< 34 weeks) and late (≥ 34 weeks).^2^ Early-onset PE is considered a fetal disorder (placental dysfunction). By contrast, late-onset PE is considered a maternal disorder (underlying maternal constitutional disorder with a normal placenta).^2^ Thus, it is hypothesized that fetal and maternal risk factors contribute asymmetrically to early and late PE, respectively. Consistent with this, maternal genetic variant rs4769612 (near *FLT1*) has a stronger association with cases of late onset PE.

Clinically, PE is characterized by high blood pressure (hypertension; HTN) with a systolic blood pressure (SBP) ≥140mmHg and/or diastolic blood pressure (DBP) ≥90mmHg in addition to signs of organ damage/dysfunction, which are commonly observed through proteinuria (PRO; proteins excreted through the urine).^3^ Left untreated, PE can lead into the development of eclampsia, which is characterized by severe seizures and an increased maternal and fetal morbidity and mortality. PE increases the risk that the fetus will be born preterm or exhibit intra-uterine growth restriction/fetal growth restriction.^4^ Despite the seriousness of PE, the only widely accepted treatment is induction of labour.^5^

PE is a complex disease, with numerous genetic and environmental factors.^6^ PE heritability has been estimated to be 54.4% maternal/42.5% fetal in Central Asians and 38.1% maternal/21.3% fetal in Europeans.^7^ The genetic variants that are associated with PE can impact on the pathophysiology in two key ways: 1) aberrant fetal-derived cytotrophoblast invasion in the uterine wall in early pregnancy can lead to failed remodeling of the maternal spiral arteries perfusing the placenta; and 2) hypoxia and/or oxidative stress to the placenta causes the release of syncytiotrophoblast-derived factors (*e.g*., receptor for vascular endothelial growth factor, VEGFR-1 (otherwise referred to as Flt-1), endothelin, and proinflammatory cytokines^8^) into the maternal circulation leading to an exaggerated inflammatory response.^9^ The release of proinflammatory cytokines contributes to the systemic inflammation associated with pregnancy, whilst Flt-1 and endothelin cause endothelial dysfunction (the likely cause of maternal HTN).^8^ Endothelial dysfunction is not unique to PE; it is also a principal pathogenic mechanism in coronary artery disease and stroke (which share HTN as a risk factor).^10^ This association has led to the study of shared genetics factors between HTN and PE.^11,12^ Similarly, sFlt-1 has also been linked with proteinuria.^13^ As such, there is likely to be an interplay of genetic factors and/or pathways that predispose to PE, HTN, and PRO.

PE involves a complex interplay between the maternal, paternal, and fetal genomes. The paternal contribution is illustrated by the finding that fathers whose mothers had PE, when carrying them, have an increased chance of the pregnancies they father being preeclamptic.^14^ Genetic variants attributed to maternal and fetal contributions to risk of developing PE have been identified by GWAS.^7,15–17^ These variants include a fetal copy-number deletion near *PSG11* and the *INHBB* locus^15,17^. Five maternally derived variants associated with PE (rs259983, rs1421085, rs4769613, rs12050029, rs149427560) also predispose individuals for HTN, consistent with the existence of a shared genetic risk between PE and HTN.^7,16^ As GWAS of PE are far smaller than similar studies in HTN (sample sizes multitudes larger than PE)^18^, it remains likely that this genetic overlap will increase as the genetics of PE are studied further.

Beyond sample size limitations, there is a need to characterize the functional roles and effects of PE-associated variants. PE-associated SNPs have typically been associated with the closest genes in the linear sequence (or the closest phenotypically relevant gene).^7,15–17,19^ However, in these regions, SNPs that associate with transcription levels of genes that are distant to the SNP locus^19^ potentially drive more of the heritable component of disease.^20^ For example, genes associated with *trans*-acting eQTLs are enriched for loss of function intolerance.^21^ Thus, a better way to annotate non-coding variants to impacted genes, protein levels, and pathways is to investigate their enrichment in control elements and assign functions via methods which attribute these variants to gene regulation.^22–24^ One approach to achieve this is to integrate information on the 3-dimensional arrangement of DNA into the functional analyses.^25^ By leveraging our knowledge of these 3D connections in relationship to loci tagged by variants associated with PE, we can better understand their role within hubs of regulation with spatially proximal genes, irrespective of their linear distance from the locus in question.

Here we have used Contextualising Developmental SNPs in 3 Dimensions (CoDeS3D) to investigate the spatially constrained eQTLs and their target genes that are associated with PE, HTN, and PRO to better understand the genetic overlap and putative biological mechanisms associated with these complex conditions. Our results identified gene candidates not previously associated with PE by published GWAS, or candidate gene studies. We then used STRING, LOEUF scores, g:Profiler, and literature queries to show that these genes converge on biological pathways and protein-protein interaction networks (PPINs) that involve PE-, HTN- and PRO-associated eQTL targeted genes.^26–29^

## Methods

### Retrieval of PE-, HTN- and PRO-associated SNPs

Unique SNPs associated (p ≤ 1 × 10^−6^) with either PE, HTN, or PRO were identified from published literature (March 2021, Search terms: “Preeclampsia GWAS”, “Hypertension GWAS”, “Proteinuria GWAS”, “Preeclampsia associated SNPs”, “Hypertension associated SNPs”, “Proteinuria associated SNPs”) and the NHGRI-EBI GWAS Catalogue (https://www.ebi.ac.uk/gwas/; downloaded March 2021; specific SNP sources found in Supplementary Table 1).^30^

SNPs were filtered to only include those with a p-value ≤ 1 × 10^−6^. SNPs were cleaned prior to use to ensure they were associated with the phenotype in question. For PE, reported traits for SNP associations included: fetal factor PE, maternal factor PE, early onset PE and late onset PE. For HTN, reported traits for all SNPs were: HTN, pulse pressure measurement, diastolic blood pressure, systolic blood pressure, early onset HTN, treatment resistant HTN, primary HTN, and mean arterial pressure. For PRO, reported traits included: PRO, albuminuria, moderate albuminuria, urinary albumin to creatine ratio, and urinary albumin excretion. For HTN, some SNPs were associated with factors considered to be out of scope for an overlap with PE, and these were therefore excluded from further analysis. These included SNPs associated with pulmonary arterial HTN, bevacizumab induced HTN, idiopathic intracranial HTN, and blood pressure response to thiazide diuretics, candesartan, calcium channel blockers and hydrochlorothiazide. For both HTN and PRO, studies that only reported associations in males were removed as, by definition, PE only occurs in females.

### eQTL and targeted gene identification and analysis

The SNP sets from each of the three conditions (PE, HTN or PRO) was independently analyzed for the existence of spatially constrained expression Quantitative Trail Loci (eQTL). For this analysis, we used Contextualizing Developmental SNPs in 3 Dimensions (CoDeS3D) to identify the regulatory impacts of genetic variants by assessing them for spatially constrained gene regulation.^29^ CoDeS3D first identifies spatial associations between sections of DNA using information from Hi-C contact data across 70 different cell lines and tissues (Supplementary Table 2).^31^ These spatial contacts were then interrogated for an association with gene expression (eQTL) from the Genotype-Tissue Expression database (GTEx), which encompasses 49 human tissues (GTEx V8).^32^ These eQTL target gene associations were divided into three classes according to the separation distance: 1) SNP and gene < 1Mb apart from one another (*cis* interactions); 2) SNP and gene > 1Mb apart (*trans*-intrachromosomal); and 3) SNP and gene on different chromosomes (*trans*-interchromosomal). Significance was corrected to control for false positives (Benjamini-Hochberg False Discovery Rate; FDR < 0.05). All SNP IDs and chromosomal positions used in this manuscript are labelled according to GRCh38.

### LOUEF score and expression variation between interaction types

LOUEF scores were obtained for all spatially constrained eQTL target genes (gnomAD database; https://gnomad.broadinstitute.org, v2.1.1^28^). A Kruskal–Wallis H test (KW) was performed to identify significant differences (p ≤ 0.05) in underlying LOUEF score distribution for each interaction type (*cis, trans*-intrachromosomal, *trans*-interchromosomal). Where significant KW results were identified, we subsequently performed a Dunn post-hoc test to determine which pairwise differences were significant (Benjamini-Hochberg FDR < 0.05).

### Mapping protein-protein interactions between PE eQTL targeted genes and directly associated HTN and PRO eQTL targeted genes

Functional protein-level connections between the spatially constrained eQTL target genes were identified using STRING (Search Tool for the Retrieval of Interacting Genes/Proteins) (https://string-db.org, version 11.0).^26^ Default settings were used, with the exception of the minimum required interaction score, which was set to 0.700 (high confidence), disconnected nodes were hidden, and active interaction sources were altered to include only text mining, experiments, databases, and co-expression. The analysis included all target genes. However, for simplification purposes, the final output for the HTN and PRO target genes was filtered to include only target genes that directly interacted with at least one PE target.

### Pathway enrichment for HTN/PRO proteins directly associated with PE proteins

Biological pathways that are impacted both by the individual genes and pathways that represent a convergence of affected genes were identified using g:Profiler (FDR < 0.05; g:GOSt – Functional profiling; https://biit.cs.ut.ee/gprofiler/gost, version *e104_eg51_p15_3922dba*)^27^ and STRING protein-protein interaction enrichment data. Only biological pathways annotated within KEGG, Reactome, and WikiPathways were considered.

### eQTL targeted gene expression in arteries, blood, and immune cell subtypes

We compared RNA expression and eQTL detection of the eQTL targeted genes within arteries (aorta, coronary and tibial) and whole blood, both identified in GTEx, whilst also identifying blood-specific spatially constrained cis-eQTLs identified within 13 different immune cell types using the Database of Immune Cell EQTLs/Expression/Epigenomics (DICE; FDR p-value ≤ 0.05; https://dice-database.org, latest build: Jan 7, 2020).^33^ All *trans*-interactions were recorded as ‘Not detected’ in DICE. Genes with TPM ≥ 0.5 are commonly considered to be expressed in a specific tissue.^34^

### Data and code availability

All datasets generated for this study are included in the Supplementary Tables and Files available in figshare with the identifier [https://doi.org/10.17608/k6.auckland.17082647.v1].

This study makes use of data generated by the GTEx Project^32^ and DICE^33^.

The CoDeS3D pipeline is available at: [https://github.com/Genome3d/codes3d-v1]. ^29^

## Results

### PE shares disease-associated SNPs, spatially constrained eQTLs and eQTL targeted genes with HTN, but not with PRO

The major diagnostic criteria for PE include elevated blood pressure (HTN) and organ damage detected through protein in the urine (PRO). Therefore, we wanted to study the potential genetic convergence of these three conditions at the SNP, gene, and protein level. SNPs associated (p ≤ 1 × 10^−6^) with PE (n= 25), HTN (n=1926), and PRO (n= 170; Supplementary Table 1) were identified. Six SNPs are associated with both PE and HTN (Figure 1A; Supplementary Table 1), and an additional six SNPs were associated with both HTN and PRO (Figure 1A; Supplementary Table 1). None of the identified SNPs were shared between PE and PRO.

**Figure 1:**
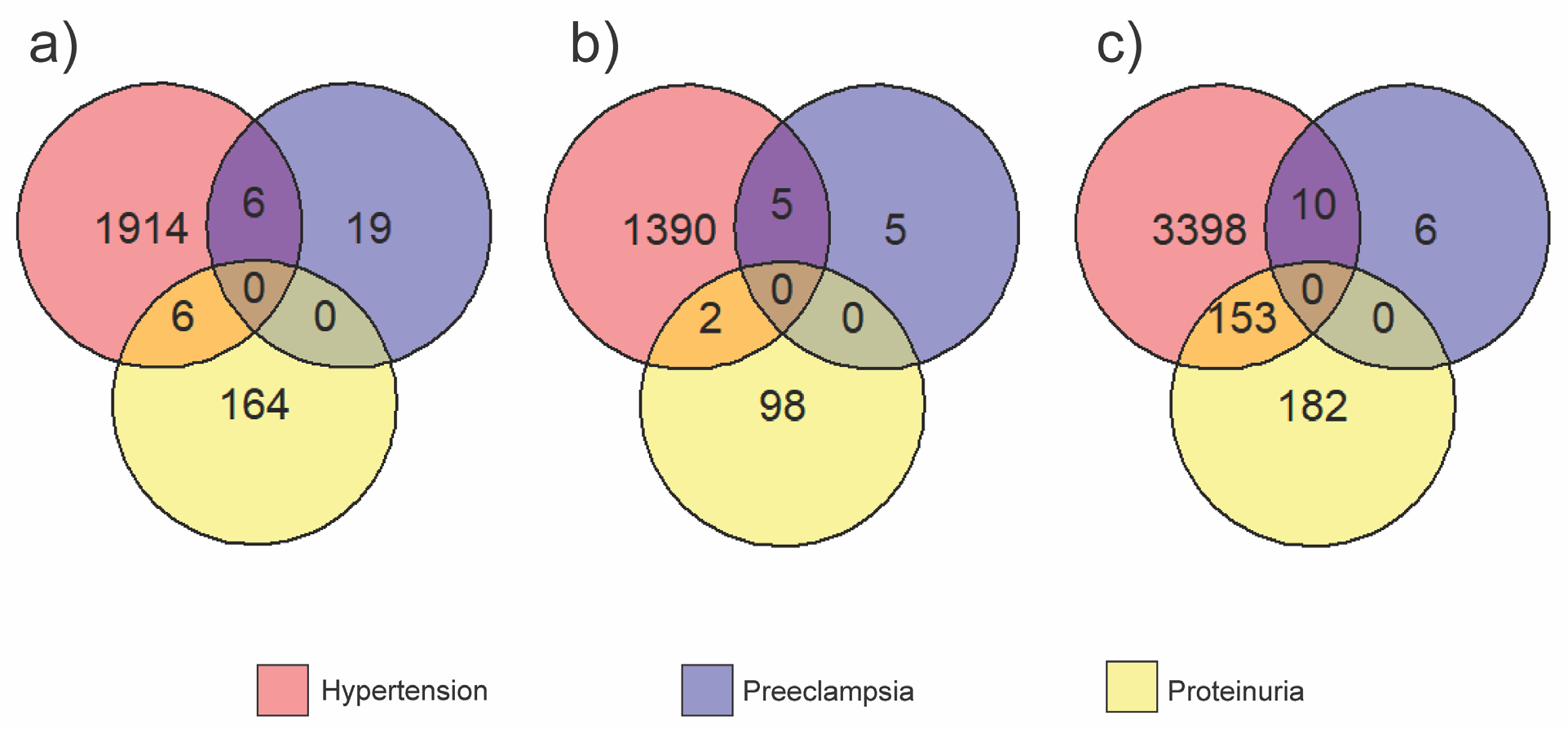
Overlaps between PE, HTN and PRO at the a) disease-associated SNP, b) eQTL and c) gene level. Numbers indicate exact counts.

Spatially constrained eQTLs were identified by screening the disease-associated SNPs through CoDeS3D (FDR q<0.05; Figure 1B; Supplementary Tables 3, 4 and 5).^29^ Of the 25 PE-associated SNPs, 15 and 4 were identified in *cis-* and *trans* eQTL-gene pairs, respectively (Table 1). Similarly, of the 1926 HTN-associated SNPs, 4402 and 1799 were identified in *cis-* and *trans* eQTL-gene pairs, respectively. Likewise, 326 *cis* eQTL-gene pairs and 125 *trans* eQTL-gene pairs were identified for the 170 PRO-associated SNPs. The number of eQTL-gene pairs can be greater than the number of disease-associated SNPs as eQTLs can have multiple gene targets in the same, or different tissues (Table 1; Supplementary Table 5). Five of the six SNPs that were shared between PE and HTN were identified as eQTLs (Figure 1A and B). Similarly, two of the six SNPs that were shared between HTN and PRO were identified as eQTLs.

**Table 1:** Summary of eQTLs associated with PE, HTN and PRO.

|  | PE | HTN | PRO |
| --- | --- | --- | --- |
| # of disease-associated SNPs<sup>1</sup> | 25 | 1926 | 170 |
| # of eQTL-gene pairs<sup>2</sup> | 41 | 28632 | 2412 |
| # of unique eQTL-gene pairs<sup>3</sup> | 19 | 6201 | 451 |
| # of unique *cis* eQTL-gene pairs<sup>4</sup> | 15 | 4402 | 326 |
| # of unique *trans* eQTL-gene pairs<sup>5</sup> | 4 | 1799 | 125 |
| # of eQTL targeted genes<sup>6</sup> | 16 | 3561 | 335 |

<sup>1</sup>Number of SNPs associated with PE, HTN or PRO from the literature. PE = Preeclampsia; HTN = Hypertension; PRO = Proteinuria. <sup>2</sup>eQTL-gene pairs were defined as a SNP that is an eQTL and its linked gene occurring in a specific tissue, therefore the same eQTL-gene pair in more than one tissue were recorded as separate eQTL-gene pairs. (FDR < 0.05). <sup>3</sup>Unique is defined as eQTL-gene pairs that occur in a minimum of one GTEx tissue (if present in more than one, the eQTL-gene pair is only counted once). <sup>4</sup>Unique eQTLs that act in *cis*; between a SNP and gene that are <1Mb apart. <sup>5</sup>Unique *trans* eQTL-gene pairs where the eQTL and gene are >1Mb apart, this includes both *trans*-intrachromosomal and *trans*-interchromosomal. <sup>6</sup>eQTL targeted genes are the genes where the expression levels are at least partially associated with the base identity of the SNP.

PE-associated eQTLs targeted 16 genes, HTN-associated eQTLs targeted 3,561 genes, and PRO-associated eQTLs targeted 335 genes (Figure 1C). Ten of the 16 PE target genes were also targeted by HTN loci (Figure 1C). Four of these 10 shared eQTL targeted genes were not due to shared SNPs, or SNPs in linkage disequilibrium, between the two conditions (Supplementary Table 8). While HTN and PRO shared 153 target genes, PE and PRO shared 0 genes.

### Mean LOEUF scores and eQTL targeted gene expression levels vary with the type of spatially constrained eQTL interaction

We used measures of mutational constraint to assign LOUEF scores (gnomAD database v2.1.1) to the spatially constrained eQTL target genes. We then tested for differences in LOUEF scores for the three interaction types (*cis, trans*-intrachromosomal, and *trans*-interchromosomal; Figure 2A, 2B and 2C, respectively; Supplementary Table 10). Mean LOUEF scores were observed to decrease as the distance between the eQTL and the target gene increased (from *cis* to *trans*-intrachromosomal to *trans*-interchromosomal; Figure 2). This is consistent with the *trans*-eQTLs being enriched for associations with genes that are intolerant to loss-of-function mutations. However, the PE results are inconclusive due to the limited number of eQTLs that were identified (Figure 2A).

**Figure 2:**
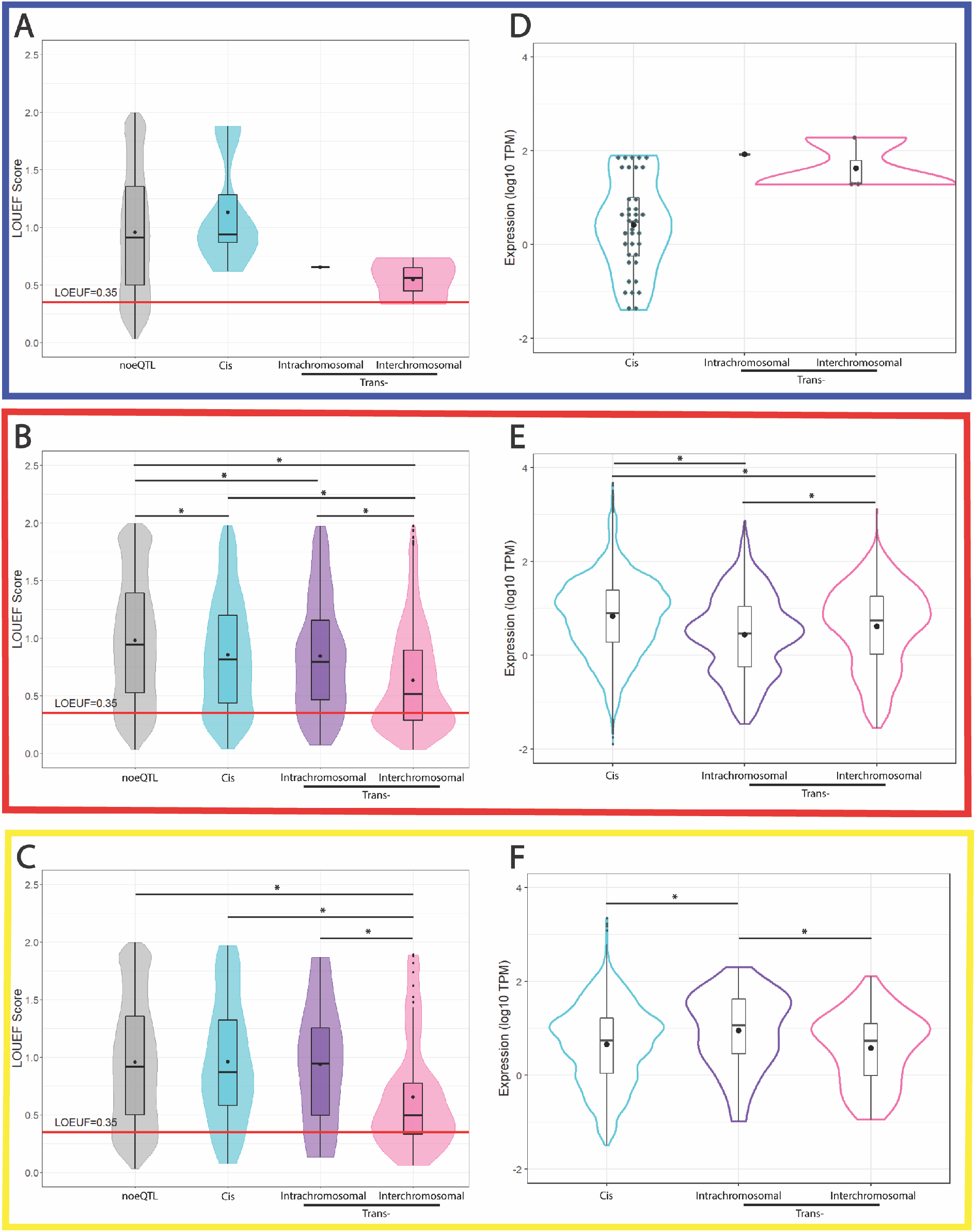
LOUEF score and expression distributions differed for genes targeted by *trans*-acting and *cis*-acting eQTLs associated with HTN and PRO. LOUEF Scores were plotted for genes according to the interaction type (*cis, trans*-intrachromosomal or *trans*-interchromosomal) for A) Preeclampsia (PE), B) hypertension (HTN), and C) proteinuria (PRO). The red line denotes the threshold between tolerant and intolerant genes (LOEUF 0.35, as defined by gnomAD^28^). The bars in the box plots denote the quartiles and median. Expression levels of the eQTL targeted genes in 49 tissues (Supplementary Tables 3, 4 and 5) were plotted for D) PE; E) HTN; and F) PRO according to eQTL interaction type (*e.g*., *cis, trans*-intrachromosomal or *trans*-interchromosomal). •, the mean LOUEF Score (A-C) or expression (D-F). *, p-value ≤ 0.05.

The mean expression levels of the eQTL target genes expression varied depending on interaction type, with significant differences for HTN (Figure 2E) and PRO (Figure 2F). Again, PE numbers prevented us testing for a significant difference (Figure 2D). Collectively, these results suggest that expression of the HTN- and PRO-associated eQTL targeted genes vary significantly depending on the associating eQTL interaction type.

### eQTL targeted gene products interact within protein-protein interaction networks (PPINs) for PE-HTN and PE-PRO

We tested for phenotypic overlap with PE (PE-HTN and PE-PRO) arising through the interaction of gene products within protein-protein interaction networks (PPIN). Convergence was observed, as clusters of interacting proteins encoded by PE-associated eQTL target genes directly interacted with proteins encoded by both HTN- and PRO-associated eQTL target genes (Figure 3A and 3B respectively). For both the PE-HTN and PE-PRO PPINs, the top 10 enriched pathways (p < 0.05) include intracellular signaling by second messengers, diseases of signal transduction by growth factor receptors and second messengers, and dopaminergic synapse (Supplementary Table 9). The large cluster within the PE-HTN PPIN contains all five PE genes that are also present in the PE-PRO PPIN (*GDNF, GSK3B, SLC2A3, FTO* and *ALDH2*; blue and purple nodes, Figure 3A and 3B). 9 of the 16 proteins within the PE-PRO PPIN were also identified within the PE-HTN PPIN. The four shared eQTL target genes between HTN and PRO (orange nodes, Figure 3) are targeted by different eQTLs (*i.e*., different SNPs targeting the same gene). Notably, the PPIN (*i.e*., biological pathway) is the first biological level at which we see an interaction between PE and PRO, following our observations that no SNPs, eQTLs, nor targeted genes are shared between these two conditions.

**Figure 3:**
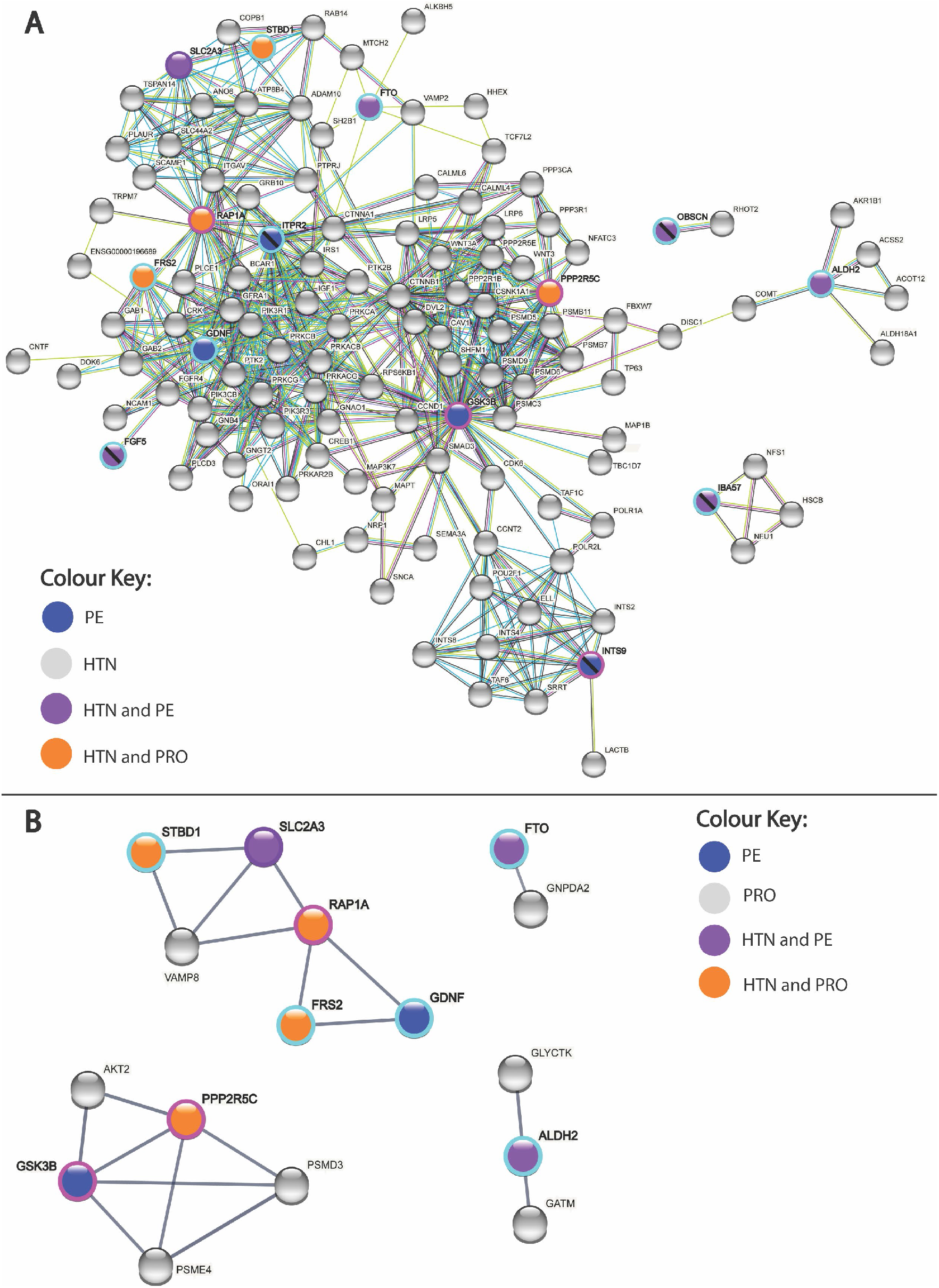
Protein-protein interaction networks occurring between eQTL targeted genes. Protein-protein interaction network for A) all PE-associated eQTL targeted genes and HTN-associated eQTL targeted genes; and B) all PE-associated eQTL targeted genes and PRO-associated eQTL targeted genes that directly interact with at least one PE-associated eQTL targeted gene. eQTL targeted genes are labelled blue (PE only), purple (PE and HTN), and orange (shared HTN and PRO). Proteins annotated with a stripe are connected to the PPIN in A) but are not B), despite being present in the dataset. Grey nodes are HTN only or PRO only eQTL targeted genes, in A and B, respectively.

### Expression and eQTL detection in artery subtypes, whole blood, and immune cells

PE is a disease of the placental arteries.^35^ Therefore, altered expression of PE-, HTN- or PRO-associated eQTL targeted genes within the blood, arteries or immune cells may be biologically relevant. From the PE-HTN PPIN (Figure 3A), three clusters were present, one large cluster containing 116 proteins (cluster PH1), cluster PH2 containing four proteins (IBA57, NFS1, NFU1 and HSCB), and cluster PH3 containing OBSCN and RHOT2. 109 of the 116 proteins in cluster PH1 were observed to have their RNA expressed (transcript levels ≥ 0.5 transcripts per million [TPM]) in at least one of the artery types (tibial, aorta and coronary) from the GTEx database (v8) (Supplementary Table 7), and 32 were targeted by a spatial eQTL detected in at least one artery subtype. In whole blood, 88 of the 116 proteins in cluster PH1 showed RNA expression (GTEx), whilst 9 were targeted by a spatial eQTL in whole blood (Supplementary Table 7). Of note, FGF5, in cluster PH1, showed no expression or eQTLs in any artery subtype or whole blood, but is targeted by an eQTL in naïve CD4+ T cells (Treg) in the DICE database (Supplementary Table 6). All four proteins in cluster PH2 were expressed in all three artery subtypes and whole blood in the GTEx database. However, only IBA57 was targeted by a spatial eQTL, in whole blood. This IBA57 eQTL was also detected in CD4^+^ T cell (Th17) in the DICE database (Supplementary Table 6). From cluster PH3, both proteins were also expressed in all artery types and whole blood in the GTEx database, but only RHOT2 was targeted by a spatial eQTL, in the tibial artery.

For the PE-PRO PPIN, four small clusters were observed: Cluster PP1 (GDNF, SLC2A3, RAP1A, STBD1, FRS2 and VAMP8), cluster PP2 (GSK3B, PPP2R5C, AKT2, PSME4, and PSMD3), cluster PP3 (ALDH2, GATM and GLYCTK) and PP4 (FTO and GNPDA2). For cluster PP1, all proteins except for GDNF were expressed in all three artery types and whole blood in the GTEx database. Yet, spatial eQTLs were only detected for STBD1 and GDNF in the tibial artery (Supplementary Table 7). For cluster PP2, all proteins are expressed in the three artery types and whole blood, but there were no spatial eQTLs targeting these genes in any artery type nor whole blood. All three genes in cluster PP3 were observed to be expressed in all three artery subtypes and whole blood in the GTEx database. Spatial eQTLs targeting all three proteins were present in at least one of the artery types, and a spatial eQTL targeting GATM and GLYCTK in whole blood. For cluster PP4, both genes were expressed in all artery types and whole blood. However, we only identified a spatial eQTL for GNPDA2 in the tibial artery.

## Discussion

The 3D conformation of the genome impacts on transcription regulation through the spatial associations that occur between regulatory elements and genes located across the genome^25^. The integration of spatial information into the assignment of putative functions to the genetic variants associated with PE-, HTN- and PRO identified a significant number of *cis*- and *trans*-acting eQTLs, with clear overlaps between PE and HTN at both the gene level and within signaling pathways. Collectively, these findings provide insights into the pathways and mechanisms that may be involved in the pathogenesis of PE and its relationships to HTN and PRO, potential targets for therapeutic intervention.

### PE and PRO converge at the pathway level but not the genetic level

PE is a hypertensive disorder with a co-occurrence of PRO in most PE cases.^3,14^ The identification of overlap at the genetic level for PE and HTN (but not PRO) suggests that the GWAS-identified PE variants have more in common with HTN than PRO. The observed overlap between PE and HTN has been identified before, where genetic factors associated with HTN have also been associated with PE.^7^ The lack of overlap at the genetic level for PE and PRO was overcome through a convergence upon shared biological pathways in the PPIN. However, the lack of any clear connection between PE and PRO at the genetic level provides support that the mechanism of PRO in PE is more as a result of the maternal HTN induced by the placenta, rather than more established kidney disease as is observed in non-pregnant populations. Thus, this provides additional support for the American Society of Obstetrics and Gynecology’s recommendation for PRO to not be a diagnostic criterion for PE.^3,36^

### Key pathways elucidated by blood- and artery-specific contributions to gene regulation in PE

The placenta is widely thought to have a significant and central role in the pathogenesis of PE. Therefore, it is important to detect whether expression changes occurred in placental tissue as a result of the disease-associated SNPs identified.^37^ However, placental (trophoblast) expression and eQTLs are not present in GTEx. Instead, we have used eQTL data for the three artery subtypes and whole blood from GTEx and verified those findings in eQTLs from the DICE database. The consideration of these tissues as significant tissues is due to the hypothetical involvement and dysfunction of these various arterial tissues in PE.^38,39^

We observed a number of proteins whose genes had altered regulation due to disease-associated eQTLs in whole blood, one of the three artery subtypes, and/or immune cell subtypes. We refer to these proteins as ‘key’ proteins due to their potential to be points of pathway disruption. From the PE-HTN PPIN, there were 41 key proteins in cluster PH1, 2 from cluster PH2 (IBA57 and NFU1), and 1 from cluster PH3 (RHOT2). From a literature search, it was found that 7 proteins in PH1 (CALML4, PIK3CB, NCAM1, GNAO1, NFATC3, FGF5, and GAB2) are altered in PE patients or are involved in certain PE related mechanisms (*e.g*., endothelial dysfunction). Decreased RNA expression of CALML4 and PIK3CB has been observed in tissue from the maternal-fetal interface of PE patients and from the decidua for early-onset PE patients respectively.^40,41^ Similarly, NCAM1 has an increased mRNA and protein expression in placentas from PE patients, and its suppression within human umbilical vein endothelial cells (HUVECs) led to increased invasiveness and migration of these cells and suppression of an MAPK signaling pathway.^42^ GNAO1 is significantly associated with early onset PE.^43^ NFATC3 has been previously observed to be involved in secretion of sFlt-1 as well as expression of proinflammatory cytokines from cytotrophoblasts.^44^ FGF5 has been previously detected as being targeted by PE- and HTN-associated variants located at the FGF5 locus.^7^ Lastly, GAB2 is upregulated in the placenta in response to systemic hypoxia and might also have a role in angiogenesis, both of which may contribute to the PE pathophysiology.^45,46^ Thus, PE-associated SNPs altering regulatory mechanisms that target these key proteins could result in a ripple effect of small alterations adding up to PE risk.

From the PE-PRO PPIN, there were seven key proteins identified: 3 (GDNF, STBD1 and VAMP8) in PP1, 3 (ALDH2, GATM and GLYCTK) in cluster PP3, and 1 (GNPDA2) in cluster PP4. Altered regulation of GATM and STBD1 has previously been observed to be associated with the PE phenotype, with one study showing increased GATM mRNA levels within PE placentae, and a different study reporting that STBD1 expression was associated with PE placentae.^47,48^ Importantly for our PE-PRO analysis, GATM is specifically involved in a creatine synthesis step that occurs specifically within the kidneys. The same kidney damage in PE that causes PRO may damage the ability of the kidneys to produce sufficient creatine, and, therefore, the increased placental expression levels of GATM could be a consequence of kidney damage having occurred.^47,49^

### Signaling pathways are the predominant pathways enriched for in the PPINs

The majority of pathways enriched for in both the PE-HTN and PE-PRO PPINs were signaling pathways. Second messengers can impact PE development, such as an increase in sensitivity to Ca^2+^ in arteries, which is influenced by numerous second messengers activating pathways such as MAPK.^38^ Additionally, two intracellular kinases (signaling pathways), PKB/Akt and MAPK/ERK, are thought to hold a strategic role in the pathogenesis of PE.^50^ This is consistent with our analysis identifying the biological pathways, ‘PIP3 activates Akt signaling’, ‘MAPK family signaling cascade’, ‘MAPK1/MAPK3 signaling’ and ‘RAF/MAP kinase cascade’ being enriched for in the PE-PRO PPIN. In addition to this, the previously discussed key gene/protein, NCAM1, was observed to be overexpressed in PE placentas, with suppression of the gene reported to suppress a particular MAPK signaling pathway, p38MAPK.^42^ Similarly, dysregulation of the Wnt/β-catenin signaling pathway has recently been identified to be a potential contributor to PE pathogenesis, which is consistent with the identification of enrichment for this pathway from the PE-HTN PPIN.^51^ Enrichment of ‘neurotrophin signaling pathways’ within our PE-PRO PPIN is supported by findings that brain derived neurotrophic factor (BDNF, a neurotrophin) has a role in placental development, angiogenesis, and the inflammatory response, as well as decreased expression in PE patients.^52^

The potential for so many pathways to be implicated in PE contributes to the complexity of understanding the disease process, as it highlights that there may be a range of different routes through which PE could develop. This suggests the possibility that PE may be able to be divided into subtypes depending on the disease variants present in the individuals, as these will cause specific genes to have altered regulation and therefore specific pathways that may be more likely to be disrupted. This idea of genetic subtypes within a disease has already been demonstrated through cancer genetic studies that have been able to identify subtypes of patients according to the genetic variants present, allowing tailored treatments to be created depending on the subtype, and improvements in survival rates occurring as a result of this.^53,54^

### Proposed Model for genetic influences on PE

We hypothesized a model for the genetic influences in PE pathogenesis, which involves tissue-specific gene targets as well as maternal- and fetal-derived genetic factors (Figure 4).

**Figure 4:**
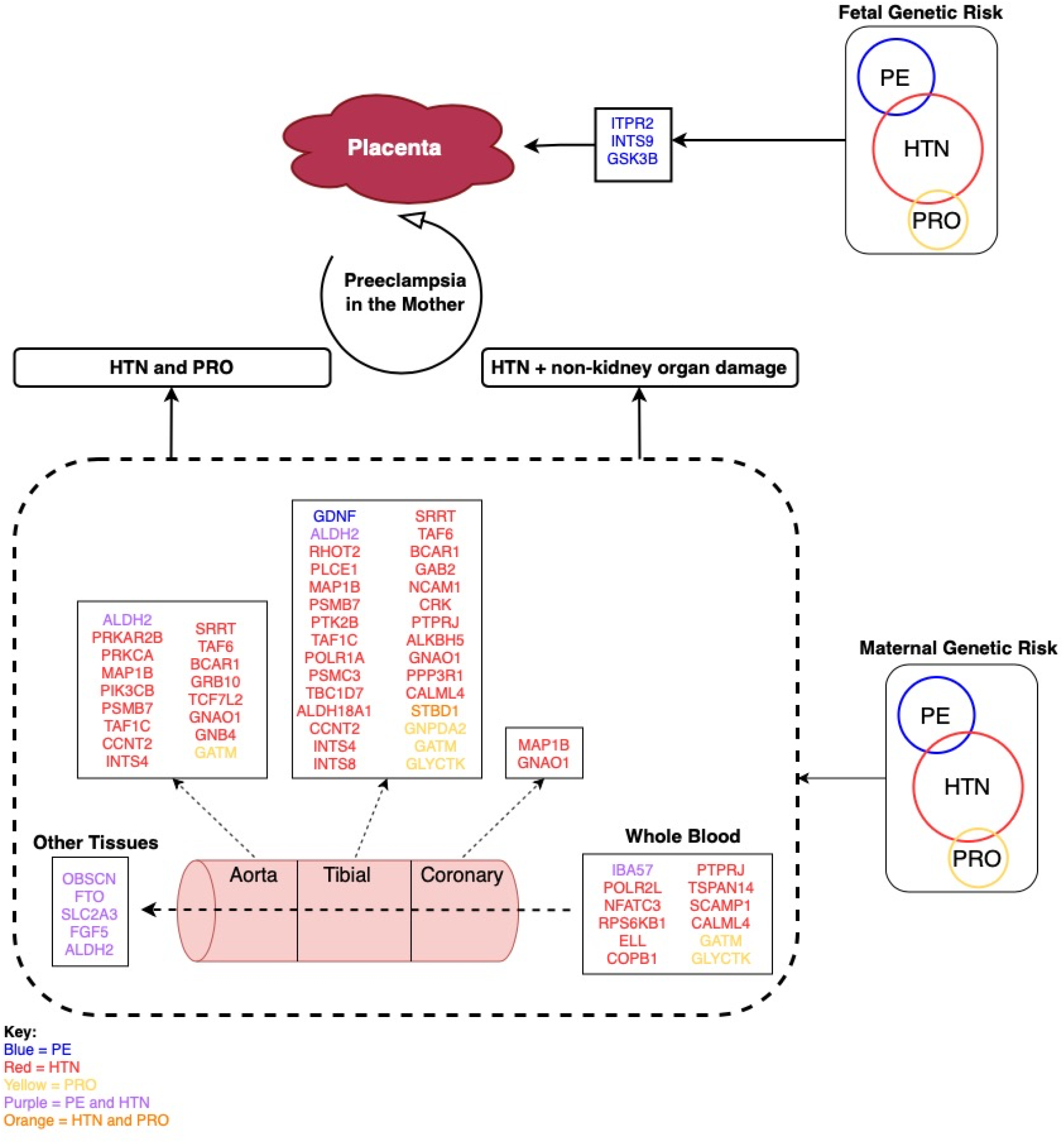
Proposed model of genetic contributions to PE development. The proteins altered in PE (blue), HTN (red), and PRO (yellow) or shared between conditions (purple or orange) contribute to the onset of PE in arterial and blood tissues. The differential patterns of genetic influence highlight the diversity of processes that may be impacted within the various tissues impacted in PE pathogenesis. Other Tissues = suprapubic not sun exposed skin, lower leg sun exposed skin, esophagus mucosa, cultured fibroblast cells, skeletal muscle, EBV transformed lymphocyte cells, and kidney cortex. Figure created on draw.io, as deployed at https://app.diagrams.net

We propose that maternal risk for PE arises through two different mechanisms: i) genes/proteins regulated by both HTN and PRO; and ii) only genes impacting HTN influence PE, with organ damage occurring as a secondary effect. For both of these mechanisms, key genes/proteins are disrupted in tissues important to both the maternal and fetal sides of the placental interface, resulting in PE in the mother. Mechanism (i) is supported by the sharing of 4 proteins between HTN and PRO at the protein and pathway level (STBD1, RAP1A, FRS2 and PPP2R5C). GSK3B, FTO, and SLC2A3 are found in both PPINs and have functions that can impact on PE pathogenesis. GSK3B is activated in response to hypoxia, an environment observed in PE patients due to inadequate placental implantation. Additionally, GSK3B, when activated, is involved in degrading a protein called GCM1, known to have a role in placental development.^55^ FTO could impact on PE through the same pathway as BMI, an independent risk factor for PE.^56,57^ SLC2A3 is a glucose transporter found within the placenta, where glucose uptake aids placental development.^58,59^ Mechanism (ii) is supported by our finding that HTN and PE form much larger networks in the PPIN, with five proteins found in the PE-HTN PPIN only (ITPR2, FGF5, INTS9, IBA57, and OBSCN). ITPR2 may contribute to PE development through its influence on vascular tone and blood pressure through Ca^2+^ release, a pathway shared by HTN and PE.^60,61^ The model also indicates the presence of key genes modified by fetal genetic risk factors, with potential roles for ITPR2 and GSK3B previously discussed as maternal factors above.

It should be noted that the exact contribution each of these genes may have to the pathogenesis of PE is unknown. However, each key gene is thought to contribute fractionally to its development, and it is a combinatorial effect of a number of them that results in PE pathology.

### Limitations of the GTEx tissue data and future directions

The predominate limitation of using GTEx data is that the tissue donor characteristics are not a reflection of the characteristics of individuals in which PE occurs. GTEx samples were isolated from a cohort of male (67.1%) and female (32.9%) tissue donors, none of whom were pregnant (GTEx database, V8 Donor info). Numerous studies have demonstrated that age has an impact on gene expression, suggesting that data sourced from individuals who are beyond child-bearing age is not ideal when looking at a condition that only occurs in females of reproductive age.^62^ Therefore, the extrapolation of these results to PE pathogenesis requires further experimental and longitudinal sampling during pregnancy. It would be advantageous to replicate these gene regulation studies in placental tissue subsets (*e.g*., trophoblasts) across varying gestational ages (*e.g*., first vs third trimester), as this would allow identification of eQTLs that may be only occurring in the placenta at specific developmental timepoints. This would help identify pathways specific to certain PE subtypes (*e.g*., early vs late PE), built on the basis that these subtypes may have different development pathways.

Notwithstanding these limitations, we were able to demonstrate tissue-specificity of our disease-associated eQTLs and were able to highlight potential differences in contributions from both the maternal and fetal genomes.

### Concluding remarks

In conclusion, we were able to identify a significant number of eQTLs associated with PE, HTN and PRO and the biological pathways they affect. The identification of overlap between HTN and PE at the genetic and protein levels distinguishes HTN from PRO in terms of their relationships with PE.

## Data Availability

All data produced in the present work are contained in the manuscript or online at https://doi.org/10.17608/k6.auckland.17082647.v1

## Supplementary Tables

**Supplementary Table 1: SNP Source Excel File**

**Supplementary Table 2: Hi-C Dataset Excel File**

**Supplementary Table 3: CoDeS3D Output PE**

**Supplementary Table 4: CoDeS3D Output HTN**

**Supplementary Table 5: CoDeS3D Output PRO**

**Supplementary Table 6: DICE Data**

**Supplementary Table 7: GTEx Artery Data**

**Supplementary Table 8: LD Table of SNPs linked to shared PE and HTN eQTL targeted genes**

**Supplementary Table 9: PPIN Enrichment Tables**

**Supplementary Table 10: Kruskal Wallis and Dunn post-hoc Test Results**

## Acknowledgements

JOS was funded by donations from the Dines Family trust. WS was supported by a postdoctoral fellowship from the Auckland Medical Research Foundation (grant ID 1320002) and a Royal Society of New Zealand Marsden Grant (20-UOA-002). This work contains data from the Genotype-Tissue Expression (GTEx) Project, which was supported by the Common Fund of the Office of the Director of the National Institutes of Health, and by NCI, NHGRI, NHLBI, NIDA, NIMH, and NINDS.

